# Characterizing adolescent-reported experiences of food insecurity in the ten Canadian provinces in 2019, 2020, and 2021: A cross-sectional analysis

**DOI:** 10.64898/2026.07.09.26357674

**Authors:** Alexandra Pepetone, Edward A. Frongillo, Lana Vanderlee, Warren Dodd, Michael P. Wallace, Joel A. Dubin, Kevin W. Dodd, David Hammond, Sharon I. Kirkpatrick

## Abstract

**Objectives:** Estimate the prevalence and sociodemographic correlates of adolescent-reported food insecurity experiences from 2019-2021.

**Methods:** Repeat cross-sectional data were collected in November-December 2019, 2020, and 2021 from adolescents aged 10-17 years living in the ten Canadian provinces (n = 11,057). The prevalence of ten items and five food insecurity subconstructs based on the 10-item Child Food Insecurity Experiences Scale was estimated. Weighted multinomial logistic regression assessed associations between sociodemographic characteristics and food insecurity experiences as a four-level (*no*, *few*, *several*, or *many* experiences) variable.

**Results:** Across 2019-2021 among adolescents, the prevalence of *worrying about food scarcity* ranged between 18.4%-22.5%, *worrying about parental/guardian ability to get food* ranged between 22.8%-26.9%, and *not being able to get the food they wanted* ranged between 23.5%-26.1%. Close to or above one in four adolescents affirmed the *uncertainty* (range: 26.9%-29.9%) and *compromised diet quality or preferences* (range: 23.5%-26.1%) subconstructs. In 2021, adolescents identifying as Black had a higher relative risk ratio of *few* food insecurity experiences (adjusted relative risk ratio (ARRR): 2.04 [95% CI: 1.20, 3.47], p-value: <0.01) and adolescents identifying as Indigenous had a higher relative risk ratio of *several* food insecurity experiences (2.38 [1.10, 5.15], p-value 0.03) compared to adolescents identifying as White. The relative risk ratio of having *few*, *several*, or *many* food insecurity experiences also differed by age, sex-at-birth, perceived income adequacy, and region.

**Conclusion:** The type and number of experiences reported underscores the value of directly measuring food insecurity. Interventions to mitigate food insecurity’s adverse consequences are warranted.

## Introduction

Household food insecurity is the inability to access adequate food due to financial constraints (Li et al., 2023). At the household level, manifestations of food insecurity range from worrying about running out of food, to not being able to afford balanced meals, to going a whole day without eating (Li et al., 2023). Measurement of household food insecurity includes indirect assessment of food insecurity experiences among individuals under 18 years of age. Based on 2021 Canadian Income Survey data collected across the provinces from January to June 2022, 24% of individuals under the age of 18 years lived in food-insecure households (Li et al., 2023). For this Survey, the Household Food Security Survey Module (HFSSM) was administered to household members aged 15 years and older (Li et al., 2023) such that information for children under 15 years was reported by another household member. Adolescents aged 12 to 17 years identifying as Black or Indigenous, living in households with a single caregiver, or living in households drawing upon social assistance were more likely to live in food-insecure households as reported using the HFSSM than individuals identifying as White, living in households with two parents, or living in households drawing upon employment income (Liu et al., 2023).

In many surveys and studies, food insecurity experiences among children and adolescents are reported by adults (Fram et al., 2011). For example, the HFSSM, typically administered to adults, includes eight questions about the experiences of children in the household (Government of Canada, 2020). Early research suggested that food insecurity is a managed process whereby adults compromise their food intake, protecting children until circumstances are dire and it is no longer possible to shield the children (Radimer et al., 1992). It was, therefore, determined that adults had sufficient knowledge to report food insecurity experiences among children and adolescents living in the same household (Radimer et al., 1992). More recent research indicates that adults can under-estimate the prevalence food insecurity experiences and miss associations between food insecurity and outcomes, e.g., poor diet quality and school absenteeism, among children and adolescents (Bernal et al., 2014, 2016; Carlos Chavez et al., 2017; Fram et al., 2013; Frongillo et al., 2019). Food insecurity among adults is also experienced differently than among children and adolescents. Among adults, food insecurity is characterized by compromises in food quality and quantity, as well as psychological and social effects (Radimer et al., 1992). Children and adolescents have been found to experience cognitive, emotional, and physical awareness of food insecurity and take responsibility for limited food resources by engaging in food saving strategies initiated by their caregiver, initiating their own strategies to stretch limited resources, and undertaking activities to increase access to food (Fram et al., 2011).

The 10-item Child Food Insecurity Experiences Scale (CFIES) was developed to capture experiences of food insecurity directly from children and adolescents (Frongillo et al., 2022). The CFIES captures the frequency of occurrences of food insecurity experiences unique to children and adolescents, including worrying about their parents’ ability to access food and feeling shame. CFIES items map to five subconstructs of food insecurity experiences: *uncertainty*, *compromised diet quality or preferences*, *eating less*, *going hungry*, and *emotional awareness* (Frongillo et al., 2022). Understanding the specific items and subconstructs of food insecurity experienced by adolescents is critical for accurately quantifying the scale and nature of this problem. Direct measurement can also support the evaluation of whether policy responses advocated to address household-level food insecurity are adequately and effectively ameliorating adolescent food insecurity experiences.

Prior work has drawn upon data using the CFIES—administered to adolescents aged 10 to 17 years and living in the Canadian provinces—to examine whether the prevalence of adolescent-reported food insecurity changed from 2019 to 2020 (Pepetone et al., 2023). To more fully characterize adolescent food insecurity experiences over time, we estimated the frequency of adolescent-reported food insecurity experiences, and its subconstructs, in 2019, 2020, and 2021. We also examined associations between age, sex-at-birth, racial-ethnic identity, income adequacy, and region and adolescent-reported food insecurity. The focus was to describe experiences of food insecurity and their correlates rather than to examine temporal trends.

## Methods

Cross-sectional data were collected from adolescents aged 10 to 17 years living in Canada in November to December in 2019, 2020, and 2021 as part of the International Food Policy Study (Hammond et al., 2022). Respondents were recruited through parents/guardians enrolled in the Nielsen Consumer Insights Global Panel and their partners’ panels. Email invitations with unique survey links were sent to adult panelists living in Canada. Those who confirmed they had an adolescent living in their household were asked for permission for their adolescent to complete a web-based survey (one adolescent per household was invited). After confirming individuals were within the eligible age range, potential adolescent respondents were provided with information about the study and asked to provide assent. Surveys were conducted in English or French. Median survey completion time was 23 minutes in 2019, 24 minutes in 2020, and 21 minutes in 2021. The adolescent’s parent/guardian received remuneration in accordance with their panel’s incentive structure (e.g., points-based or monetary rewards, chances to win prizes). The study was reviewed by and received ethics clearance through a University of Waterloo Research Ethics Board (REB# 41477).

In 2019, 3,825 (0.9% response rate) were surveyed, with 4,000 adolescents (5.9% response rate) surveyed in 2020, and 3,652 (15.7% response rate) surveyed in 2021. Respondent data were excluded (2019: n = 143; 2020: n = 105; 2021: n = 153) because of missing or ineligible region information (i.e., stating a region outside of Canada), living in a region with inadequate sample size to reliably construct sample weights (i.e., in the territories), invalid response to a data quality question, survey completion time under ten minutes in 2019 and 2020 and under eight minutes in 2021, and/or multiple invalid responses to open-ended measures. After excluding respondents with missing data on food security experiences (2019: n = 8; 2020: n = 5; 2021: n = 6), the final analytic samples included 3,674 respondents in 2019, 3,890 respondents in 2020, and 3,493 respondents in 2021 living in the ten Canadian provinces.

### Measures

Experiences of food insecurity in the previous 12 months were assessed in November to December of each year using the 10-item CFIES (**Table 1**). Acceptable criterion validity of the CFIES relative to other markers of food insecurity has been demonstrated among individuals aged 5 to 18 years in 13 countries (Frongillo et al., 2022). Across 15 surveys incorporating the CFIES, cross-contextual equivalence was demonstrated using the alignment method (multiple-group confirmatory factor analysis), with 83% equivalence (Frongillo et al., 2022). The Cronbach’s alpha reliability coefficient ranged from 0.88 to 0.94 and was 0.92 in Canada (Frongillo et al., 2022).

**Table 1:** Food insecurity items and corresponding subconstructs among adolescents.

| **Items** | **Subconstructs** |
| --- | --- |
| *Worry about food scarcity*<br>*Worry about parental/guardian ability to get food* | *Uncertainty* |
| *Not able to get food wanted due to lack of money* | *Compromised diet quality or preferences* |
| *Cut size of meal due to lack of food*<br>*Skipped meal due to lack of food* | *Eating less* |
| *Hungry and unable to eat due to lack of food*<br>*Tired or weak due to lack of food* | *Going hungry* |
| *Shame about lack of food*<br>*Sad or mad about lack of food*<br>*Shame about methods used to get food* | *Emotional awareness* |

Each item was scored from zero to two, with a score of 0 indicating never having the specified experience in the previous 12 months, a 1 indicating one or two times, and a 2 indicating many times. Scores were summed and a four-level ordinal variable indicating *no* food insecurity experiences (score: 0), *few* experiences (score: 1 to 6), *several* experiences (score: 7 to 10), and *many* experiences (score: 11 to 20) was created (Frongillo et al., 2022). Dichotomous variables were created for the items and subconstructs. For each item, a scored value of 1 or 2 indicated having the food insecurity experience specified while a scored value of 0 indicated not having that experience in the last 12 months. For subconstructs (*uncertainty*, *compromised diet quality or preferences*, *eating less*, *going hungry*, and *emotional awareness)*, a score of 1 or more indicated affirming at least one experience and a score of 0 indicated no affirmation of experiences related to the subconstruct.

Potential correlates of food insecurity were based on prior research (Hutchinson & Tarasuk, 2022; Kirkpatrick & Tarasuk, 2008; Li et al., 2023; Liu et al., 2023). Respondents reported their age from 10 to 17 years and their sex-at-birth as female or male. Racial-ethnic identity was ascertained using ten multi-select response options, which included White, East/Southeast Asian, South Asian, Black, Indigenous, Latino, Middle Eastern, the option for respondents to specify their own identity, as well as don’t know and refuse to answer. To mitigate small cell counts, responses were consolidated into eight categories including Black only, East/Southeast Asian only, Indigenous only, Latino only, Middle Eastern only, South Asian only, White only, Mixed/other/not stated/missing. Perceived income adequacy was ascertained by asking adolescents, “Does your family have enough money to pay for things your family needs?” (Acton et al., 2025). Based on the distribution of responses, a three-category variable indicating not enough money (not enough and barely enough money), enough money (enough and more than enough), and not stated (don’t know and refuse to answer) was created. Responses for region were Atlantic provinces (includes New Brunswick, Newfoundland and Labrador, Nova Scotia, Prince Edward Island), Québec, Ontario, Prairie provinces (includes Alberta, Manitoba, Saskatchewan), and British Columbia.

### Statistical analysis

Data were weighted with post-stratification sample weights constructed using a raking algorithm and census data on age, sex-at-birth, and region (Hammond et al., 2022). Analyses were conducted using SAS, version 9.4 (Cary, NC). SURVEY commands were applied to allow for the application of sample weights. Univariate frequency tables and means were used to characterize sociodemographic characteristics and the overall occurrence of experiences of food insecurity, as well as the prevalence of experiences of each item and subconstruct, by year. Weighted multinomial logistic regression stratified by year assessed associations between the sociodemographic characteristics—age, sex-at-birth, racial-ethnic identity, perceived income adequacy, and region—and the four-level food insecurity experiences variable. All models converged. Multinomial logistic regression was selected because it allows for *few*, *several*, and *many* food insecurity experiences to be examined independent of each other in relation to *no* food insecurity experiences. Inferences were drawn by considering the strength of evidence provided by the point estimates (i.e., relative risk ratios), p-values, and 95% confidence intervals (Muff et al., 2022).

## Results

Equal proportions of adolescents reported their sex-at-birth as female and male each year (**Table 2**). About seven-in-ten identified as White and about eight-in-ten reported their family had enough money for things they needed at all time points.

**Table 2:** Sociodemographic characteristics of adolescents aged 10-17 years living in the ten Canadian provinces in 2019, 2020, and 2021, International Food Policy Study (n = 11,057).^a^.

| **Sociodemographic characteristics** | **2019 (n = 3,674),**<br>**n (%)** | **2020 (n = 3,890),**<br>**n (%)** | **2021 (n = 3,493),**<br>**n (%)** |
| --- | --- | --- | --- |
| **Age<sup>b</sup>** | 13.5 (2.3) | 13.5 (2.3) | 13.5 (2.3) |
| **Sex-at-birth** |  |  |  |
| Female | 1755 (49.0) | 1940 (49.1) | 1593 (49.1) |
| Male | 1919 (51.0) | 1950 (50.9) | 1900 (50.9) |
| **Racial-ethnic identity** |  |  |  |
| Black only | 93 (2.6) | 108 (2.8) | 90 (2.7) |
| East/Southeast Asian only | 250 (7.5) | 340 (9.2) | 268 (8.8) |
| Indigenous only | 73 (2.1) | 71 (2.0) | 122 (3.6) |
| Latino only | 52 (1.4) | 48 (1.3) | 45 (1.5) |
| Middle Eastern only | 53 (1.5) | 53 (1.4) | 50 (1.4) |
| South Asian only | 141 (4.2) | 188 (5.1) | 147 (4.5) |
| White only | 2735 (72.8) | 2745 (69.3) | 2505 (69.4) |
| Mixed/other/not stated/missing<sup>c</sup> | 277 (7.9) | 337 (9.0) | 266 (8.1) |
| **Perceived income adequacy** |  |  |  |
| Not enough money | 632 (17.3) | 686 (17.9) | 617 (18.1) |
| Enough money | 2998 (81.4) | 3146 (80.5) | 2839 (80.8) |
| Not stated | 44 (1.3) | 58 (1.6) | 37 (1.0) |
| **Region** |  |  |  |
| Atlantic provinces | 311 (6.1) | 321 (6.1) | 259 (6.0) |
| Québec | 973 (21.3) | 966 (21.6) | 982 (21.9) |
| Ontario | 1366 (39.4) | 1553 (39.0) | 1352 (38.7) |
| Prairie provinces | 658 (20.8) | 638 (21.0) | 596 (21.1) |
| British Columbia | 366 (12.4) | 412 (12.3) | 304 (12.3) |

<sup>a</sup>Percentages may not sum to 100% owing to rounding. Cell counts are unweighted and may not align with the percentages which are weighted.
<sup>b</sup>Mean and standard deviation.
<sup>c</sup>The mixed/other/not stated/missing category can be broken down in the following manner: mixed racial-ethnic identity: 170 in 2019, 228 in 2020, 184 in 2021; other: 41 in 2019, 53 in 2020, 38 in 2021; don't know: 28 in 2019, 25 in 2020, 25 in 2021; refused to answer: 38 in 2019, 31 in 2020, 19 in 2021.

In 2019, 36.4% of adolescents reported one or more experiences of food insecurity in the past year (**Table 3**). Similar prevalences were observed in 2020 (35.0%) and 2021 (37.0%) (**Table 3**). Across all years, close to or more than one in five to one in four adolescents reported *worrying about food scarcity*, *worrying about parental/guardian ability to get food*, and *not being able to get the food they wanted* (**Table 3**). The remaining items were affirmed by 6% to 16% of adolescents each year. Close to or more than one in four adolescents had experiences of food insecurity related to the *uncertainty* and *compromised diet quality or preferences* subconstructs (**Table 3**). The *eating less*, *going hungry*, and *emotional awareness* subconstructs were experienced by 9% to 20% of adolescents each year.

**Table 3:** Estimated prevalence of adolescents aged 10-17 years living in the ten Canadian provinces in 2019, 2020, and 2021 who experienced food insecurity, who affirmed each item (score of 1 or 2) in the Child Food Insecurity Experiences Scale, and who had experiences related to each subconstruct of food insecurity in the past 12 months, International Food Policy Study (n = 11,057).

| **Conceptualizations of food insecurity** | **2019 (n = 3,674),**<br>**% (95% CI)** | **2020 (n = 3,890),**<br>**% (95% CI)** | **2021 (n = 3,493),**<br>**% (95% CI)** |
| --- | --- | --- | --- |
| **Number of food insecurity experiences** |  |  |  |
| *No experiences* | 63.6 (62.0, 65.2) | 65.0 (63.4, 66.5) | 63.0 (61.3, 64.6) |
| *Few experiences* | 27.3 (25.9, 28.8) | 26.7 (25.3, 28.2) | 23.7 (22.2, 25.2) |
| *Several experiences* | 4.6 (3.9, 5.3) | 4.2 (3.6, 4.9) | 5.0 (4.2, 5.7) |
| *Many experiences* | 4.4 (3.7, 5.1) | 4.1 (3.4, 4.7) | 8.3 (7.4, 9.3) |
| **Food insecurity items** |  |  |  |
| *Worry about food scarcity* | 18.8 (17.5, 20.1) | 18.4 (17.2, 19.7) | 22.5 (21.1, 23.9) |
| *Worry about parental/guardian ability to get food* | 22.8 (21.4, 24.2) | 25.1 (23.7, 26.5) | 26.9 (25.3, 28.4) |
| *Not able to get food wanted due to lack of money* | 26.1 (24.6, 27.6) | 23.5 (22.1, 24.8) | 26.0 (24.5, 27.5) |
| *Cut size of meal due to lack of food* | 11.4 (10.4, 12.5) | 9.8 (8.9, 10.8) | 15.8 (14.5, 17.0) |
| *Hungry and unable to eat due to lack of food* | 9.0 (8.1, 10.0) | 7.4 (6.5, 8.2) | 12.8 (11.7, 14.0) |
| *Skipped meal due to lack of food* | 8.3 (7.3, 9.2) | 7.5 (6.6, 8.3) | 12.9 (11.7, 14.0) |
| *Tired or weak due to lack of food* | 7.2 (6.3, 8.1) | 6.3 (5.5, 7.1) | 11.8 (10.7, 12.8) |
| *Shame about lack of food* | 8.9 (8.0, 9.9) | 8.0 (7.1, 8.9) | 13.6 (12.4, 14.8) |
| *Sad or mad about lack of food* | 11.5 (10.4, 12.5) | 10.6 (9.6, 11.6) | 16.2 (14.9, 17.4) |
| *Shame about methods used to get food* | 8.0 (7.1, 8.9) | 7.6 (6.8, 8.5) | 13.5 (12.4, 14.7) |
| **Subconstructs of food insecurity** |  |  |  |
| *Uncertainty* | 26.9 (25.4, 28.4) | 27.9 (26.5, 29.4) | 29.9 (28.4, 31.5) |
| *Compromised diet quality or preferences* | 26.1 (24.6, 27.6) | 23.5 (22.1, 24.8) | 26.0 (24.5, 27.5) |
| *Eating less* | 13.4 (12.3, 14.6) | 11.6 (10.5, 12.6) | 18.0 (16.6, 19.3) |
| *Going hungry* | 10.7 (9.6, 11.7) | 8.9 (8.0, 9.8) | 15.1 (13.9, 16.3) |
| *Emotional awareness* | 13.9 (12.7, 15.0) | 12.5 (11.4, 13.5) | 19.5 (18.2, 20.9) |

In most instances, there was little evidence of differences in the relative risk ratio of *few*, *several*, and *many* compared to *no* food insecurity experiences by age in 2019, 2020, and 2021 (**Tables 4 to 6**). Females had a lower relative risk ratio of *several* or *many* food insecurity experiences compared to males in 2019 (*many* -adjusted relative risk ratio (ARRR): 0.67 [95% CI: 0.47, 0.95], p-value: 0.03), 2020 (*several* -0.61 [0.43, 0.88], p-value: <0.01), and 2021 (*several* -0.54 [0.38, 0.76], p-value: <0.001; *many* -0.69 [0.52, 0.91], p-value: <0.01) (**Tables 4** to **6**).

**Table 4:** Adjusted relative risk ratio of *few*, *several*, and *many* compared to *no* food insecurity experiences in the past 12 months in relation to sociodemographic characteristics among adolescents aged 10-17 years living in the ten Canadian provinces in 2019, International Food Policy Study (n = 3,674).

| Experiences of food insecurity | Few |  | Several |  | Many |  |
| --- | --- | --- | --- | --- | --- | --- |
|  | Adjusted relative risk ratio (95% CI) | p-value | Adjusted relative risk ratio (95% CI) | p-value | Adjusted relative risk ratio (95% CI) | p-value |
| **Age** | 0.97 (0.94, 1.01) | 0.10 | 1.00 (0.93, 1.08) | 0.93 | 1.02 (0.94, 1.11) | 0.64 |
| **Sex-at-birth** (reference = Male) |  |  |  |  |  |  |
| Female | 1.09 (0.93, 1.29) | 0.28 | 1.15 (0.81, 1.62) | 0.44 | 0.67 (0.47, 0.95) | 0.03 |
| **Racial-ethnic identity** (reference = White only) |  |  |  |  |  |  |
| Black only | 1.17 (0.69, 1.99) | 0.57 | 1.54 (0.61, 3.86) | 0.36 | 2.68 (1.25, 5.72) | 0.01 |
| East/Southeast Asian only | 0.65 (0.46, 0.94) | 0.02 | 0.79 (0.37, 1.72) | 0.56 | 0.46 (0.17, 1.21) | 0.11 |
| Indigenous only | 2.09 (1.17, 3.71) | 0.01 | 1.07 (0.35, 3.27) | 0.91 | 1.51 (0.54, 4.25) | 0.44 |
| Latino only | 1.26 (0.63, 2.52) | 0.51 | 1.40 (0.28, 6.92) | 0.68 | 1.74 (0.49, 6.25) | 0.39 |
| Middle Eastern only | 0.69 (0.31, 1.54) | 0.37 | 1.63 (0.35, 7.66) | 0.53 | 0.51 (0.07, 3.58) | 0.50 |
| South Asian only | 1.25 (0.81, 1.95) | 0.32 | 2.42 (1.13, 5.18) | 0.02 | 1.27 (0.50, 3.25) | 0.61 |
| Mixed/other/not stated/missing | 1.10 (0.81, 1.49) | 0.55 | 1.00 (0.54, 1.83) | 0.99 | 0.97 (0.51, 1.84) | 0.93 |
| **Perceived income adequacy** (reference = Enough money) |  |  |  |  |  |  |
| Not enough money | 8.84 (6.96, 11.23) | <0.0001 | 31.97 (21.98, 46.50) | <0.0001 | 26.33 (18.10, 38.31) | <0.0001 |
| Not stated | 3.16 (1.64, 6.07) | <0.001 | 2.71 (0.58, 12.61) | 0.20 | 2.73 (0.35, 21.17) | 0.34 |
| **Region** (reference = Ontario) |  |  |  |  |  |  |
| Atlantic provinces | 1.13 (0.83, 1.52) | 0.44 | 0.88 (0.46, 1.66) | 0.69 | 1.33 (0.71, 2.50) | 0.38 |
| Québec | 0.73 (0.59, 0.91) | <0.01 | 0.90 (0.58, 1.39) | 0.63 | 0.81 (0.49, 1.33) | 0.40 |
| Prairie provinces | 1.13 (0.89, 1.42) | 0.31 | 0.97 (0.60, 1.57) | 0.89 | 1.60 (1.00, 2.58) | 0.05 |
| British Columbia | 1.42 (1.07, 1.87) | 0.01 | 1.50 (0.85, 2.65) | 0.16 | 2.35 (1.32, 4.17) | <0.01 |

In 2019, adolescents identifying as Black, Indigenous, and South Asian compared to those identifying as White had a higher relative risk ratio of *few*, *several* and *many* compared to *no* food insecurity experiences (**Table 4**). There was moderate evidence that adolescents identifying as East/Southeast Asian had a lower relative risk ratio of *few* (0.65 [0.46, 0.94], p-value: 0.02) compared to *no* food insecurity experiences relative to those identifying as White. Adolescents identifying as Black, East/Southeast Asian, Indigenous, Latino, and South Asian compared to White had a higher relative risk ratio of *few*, *several*, and *many* food insecurity experiences in 2020 (**Table 5**). In 2021, adolescents identifying as Black had a higher relative risk ratio of *few* (2.04 [1.20, 3.47], p-value: <0.01) food insecurity experiences and adolescents identifying as Indigenous had a higher relative risk ratio of *several* (2.38 [1.10, 5.15], p-value 0.03) food insecurity experiences compared to adolescents identifying as White (**Table 6**). Adolescents identifying as East/Southeast Asian relative to those identifying as White had a lower relative risk ratio of *many* (0.27 [0.13, 0.57], p-value: <0.001) food insecurity experiences in 2021.

**Table 5:** Adjusted relative risk ratio of *few*, *several*, and *many* compared to *no* food insecurity experiences in the past 12 months in relation to sociodemographic characteristics among adolescents aged 10-17 years living in the ten Canadian provinces in 2020, International Food Policy Study (n = 3,890).

| Experiences of food insecurity | Few |  | Several |  | Many |  |
| --- | --- | --- | --- | --- | --- | --- |
|  | Adjusted relative risk ratio (95% CI) | p-value | Adjusted relative risk ratio (95% CI) | p-value | Adjusted relative risk ratio (95% CI) | p-value |
| **Age** | 0.99 (0.95, 1.03) | 0.57 | 1.02 (0.95, 1.10) | 0.53 | 1.10 (1.02, 1.19) | 0.01 |
| **Sex-at-birth** (reference = Male) |  |  |  |  |  |  |
| Female | 1.11 (0.95, 1.31) | 0.20 | 0.61 (0.43, 0.88) | <0.01 | 0.84 (0.59, 1.20) | 0.33 |
| **Racial-ethnic identity** (reference = White only) |  |  |  |  |  |  |
| Black only | 2.10 (1.33, 3.31) | <0.01 | 2.53 (0.85, 7.55) | 0.10 | 5.10 (2.53, 10.28) | <0.0001 |
| East/Southeast Asian only | 0.94 (0.70, 1.26) | 0.67 | 3.18 (1.88, 5.38) | <0.0001 | 0.69 (0.33, 1.47) | 0.33 |
| Indigenous only | 1.86 (1.04, 3.31) | 0.04 | 3.39 (1.18, 9.77) | 0.02 | 2.96 (1.16, 7.59) | 0.02 |
| Latino only | 3.08 (1.43, 6.65) | <0.01 | 12.39 (4.23, 36.32) | <0.0001 | 0.88 (0.11, 7.41) | 0.91 |
| Middle Eastern only | 1.09 (0.54, 2.17) | 0.82 | 3.12 (1.00, 9.76) | 0.05 | 0.82 (0.15, 4.45) | 0.82 |
| South Asian only | 1.30 (0.89, 1.89) | 0.18 | 2.91 (1.45, 5.83) | <0.01 | 2.37 (1.20, 4.69) | 0.01 |
| Mixed/other/not stated/missing | 1.23 (0.93, 1.63) | 0.15 | 2.21 (1.27, 3.86) | <0.01 | 1.01 (0.54, 1.91) | 0.96 |
| **Perceived income adequacy** (reference = Enough money) |  |  |  |  |  |  |
| Not enough money | 8.29 (6.62, 10.39) | <0.0001 | 27.92 (18.96, 41.13) | <0.0001 | 37.26 (25.00, 55.55) | <0.0001 |
| Not stated | 1.60 (0.86, 2.95) | 0.14 | 3.72 (1.18, 11.78) | 0.03 | 2.43 (0.67, 8.80) | 0.18 |
| **Region** (reference = Ontario) |  |  |  |  |  |  |
| Atlantic provinces | 0.77 (0.57, 1.05) | 0.10 | 1.15 (0.59, 2.24) | 0.67 | 0.67 (0.33, 1.35) | 0.26 |
| Québec | 0.55 (0.44, 0.69) | <0.0001 | 0.76 (0.46, 1.25) | 0.28 | 0.69 (0.42, 1.14) | 0.14 |
| Prairie provinces | 1.09 (0.87, 1.37) | 0.43 | 1.36 (0.85, 2.17) | 0.21 | 0.95 (0.57, 1.58) | 0.84 |
| British Columbia | 0.94 (0.72, 1.24) | 0.67 | 0.83 (0.46, 1.49) | 0.53 | 1.11 (0.65, 1.88) | 0.71 |

Across all years, adolescents who reported their family did not have enough money for things their family needed had a higher relative risk ratio of experiencing food insecurity compared to those reporting their family had enough money (**Tables 4** to **6**). Adolescents in Québec at all time points had a lower relative risk ratio of *few* compared to *no* experiences of food insecurity than adolescents in Ontario. There was moderate and very strong evidence, respectively, that adolescents in Québec relative to Ontario in 2021 had a lower relative risk ratio of *several* (0.48 [0.30, 0.77], p-value: <0.01) and *many* (0.33 [0.22, 0.48], p-value: <0.0001) compared to *no* experiences of food insecurity (**Table 6**).

**Table 6:** Adjusted relative risk ratio of *few*, *several*, and *many* compared to *no* food insecurity experiences in the past 12 months in relation to sociodemographic characteristics among adolescents aged 10-17 years living in the ten Canadian provinces in 2021, International Food Policy Study (n = 3,493).

| Experiences of food insecurity | Few |  | Several |  | Many |  |
| --- | --- | --- | --- | --- | --- | --- |
|  | Adjusted relative risk ratio (95% CI) | p-value | Adjusted relative risk ratio (95% CI) | p-value | Adjusted relative risk ratio (95% CI) | p-value |
| **Age** | 0.97 (0.93, 1.01) | 0.14 | 0.95 (0.88, 1.02) | 0.16 | 1.01 (0.96, 1.07) | 0.66 |
| **Sex-at-birth** (reference = Male) |  |  |  |  |  |  |
| Female | 1.12 (0.94, 1.34) | 0.21 | 0.54 (0.38, 0.76) | <0.001 | 0.69 (0.52, 0.91) | <0.01 |
| **Racial-ethnic identity** (reference = White only) |  |  |  |  |  |  |
| Black only | 2.04 (1.20, 3.47) | <0.01 | 1.87 (0.70, 5.02) | 0.21 | 1.59 (0.72, 3.52) | 0.25 |
| East/Southeast Asian only | 0.76 (0.54, 1.08) | 0.13 | 1.04 (0.57, 1.90) | 0.90 | 0.27 (0.13, 0.57) | <0.001 |
| Indigenous only | 1.13 (0.69, 1.88) | 0.63 | 2.38 (1.10, 5.15) | 0.03 | 1.73 (0.96, 3.10) | 0.07 |
| Latino only | 1.05 (0.48, 2.32) | 0.90 | 1.33 (0.40, 4.46) | 0.65 | 0.73 (0.22, 2.44) | 0.61 |
| Middle Eastern only | 1.80 (0.84, 3.87) | 0.13 | 2.85 (0.99, 8.19) | 0.05 | 1.11 (0.34, 3.59) | 0.86 |
| South Asian only | 1.16 (0.75, 1.80) | 0.50 | 1.21 (0.53, 2.76) | 0.65 | 0.33 (0.13, 0.85) | 0.02 |
| Mixed/other/not stated/missing | 0.90 (0.64, 1.27) | 0.54 | 0.79 (0.42, 1.49) | 0.47 | 0.51 (0.30, 0.86) | 0.01 |
| **Perceived income adequacy** (reference = Enough money) |  |  |  |  |  |  |
| Not enough money | 9.67 (7.54, 12.41) | <0.0001 | 21.80 (15.00, 31.68) | <0.0001 | 19.80 (14.48, 27.09) | <0.0001 |
| Not stated | 2.15 (1.01, 4.58) | <0.05 | 3.93 (1.05, 14.76) | 0.04 | Value suppressed. | N/A |
| **Region** (reference = Ontario) |  |  |  |  |  |  |
| Atlantic provinces | 1.37 (0.97, 1.93) | 0.08 | 1.43 (0.81, 2.55) | 0.22 | 0.53 (0.30, 0.92) | 0.02 |
| Québec | 0.74 (0.59, 0.94) | 0.01 | 0.48 (0.30, 0.77) | <0.01 | 0.33 (0.22, 0.48) | <0.0001 |
| Prairie provinces | 1.25 (0.97, 1.60) | 0.09 | 1.06 (0.68, 1.64) | 0.81 | 0.53 (0.36, 0.77) | <0.001 |
| British Columbia | 1.40 (1.03, 1.91) | 0.03 | 0.80 (0.43, 1.49) | 0.49 | 0.83 (0.51, 1.34) | 0.44 |

## Discussion

About one in three adolescents living in the ten Canadian provinces had experiences of food insecurity between 2019 and 2021. Many reported *worrying about food scarcity*, *worrying about parental/guardian ability to get food*, and *not being able to get the food they wanted*. These experiences were especially common among adolescents identifying as Black, East/Southeast Asian, Indigenous, Latino, and South Asian and among those who said their family did not have enough money for things they needed.

The estimated proportions of adolescents experiencing food insecurity are higher than those previously observed (Li et al., 2023; Liu et al., 2023). Differences may be due to the administration of a household-focused measure designed for adults and relying on adult reporting of children’s and adolescent’s experiences in prior studies. Although some experiences of food insecurity relevant to adolescents can be captured through adult reporting mechanisms, such as the HFSSM, they may not be accurately reported by caregivers (Fram et al., 2011; Frongillo et al., 2019, 2022; Government of Canada, 2020). For example, going hungry is an experience captured in the CFIES and the HFSSM that is difficult to detect through reporting by another person (Frongillo, 2022; Government of Canada, 2020). Prior research that used the HFSSM estimated the prevalence of children experiencing hunger in 2021 to be about 0.7% (Li et al., 2023) compared to 12.8% in 2021 reported by adolescents in the present investigation.

The CFIES captures a range of adolescent-specific experiences that are not captured by adult reporting mechanisms, such as *worry about parental/guardian ability to get food*, which was reported by close to or more than one in four adolescents in the present investigation across all years. Moreover, the structure of the CFIES (i.e., five subconstructs) focuses on the presence of different types of food insecurity experiences, in contrast to the adult-oriented HFSSM, which is unidimensional and characterizes experiences by severity (Frongillo, 2022). As a result, affirming more items on the CFIES reflects a broader range of experiences (but not necessarily more severe food insecurity) (Frongillo, 2022). Better understanding the range of experiences through the CFIES may improve our understanding of the mechanisms that underlie adverse consequences associated with food insecurity and inform avenues for tailored interventions. For example, access to information about social acceptability, i.e., feelings of shame, may help us better understand the association between food insecurity and poor mental health. Adult and adolescent reporting mechanisms are, therefore, complementary and help provide a more comprehensive understanding of food insecurity among adolescents.

Correlates of food insecurity among adolescents living in the ten Canadian provinces were similar to those observed in prior research (Hutchinson & Tarasuk, 2022; Kirkpatrick & Tarasuk, 2008; Li et al., 2023; Liu et al., 2023). Consistent findings, for instance, of the association with inadequate income, irrespective of the measure used and who reported the experiences, underscores the social and economic disadvantage that underlie food insecurity (Li et al., 2023; Liu et al., 2023). The differential association between racial-ethnic identity and food insecurity in the present investigation and prior research underscores the structural racism that portions of the population experience (Dhunna & Tarasuk, 2021; Liu et al., 2023). Addressing structural racism is, therefore, necessary to equitably ameliorate food insecurity (Dhunna & Tarasuk, 2021).

Living in Québec relative to Ontario was associated with a lower relative risk ratio of experiencing food insecurity, with the same pattern observed in prior research (Li et al., 2023). Québec’s higher level of social assistance relative to other provinces and universal childcare may support securing gainful employment (Daigneault et al., 2021; Draghici et al., 2023). Relying on social assistance as a main source of income compared to employment income is associated with food insecurity (Liu et al., 2023). Social assistance in Québec is indexed to inflation, potentially supporting mobility into gainful employment opportunities that mitigate food insecurity experiences (Laidley & Tabbara, 2023). Responses to the COVID-19 pandemic in 2020 and 2021 likely also contributed to differences in food insecurity across regions. For example, eligibility for provincial and territorial social assistance was, in several instances, altered by the receipt of the Canada Emergency Response Benefit, a national level government policy response during the pandemic (Tweddle & Stapleton, 2020). These changes influenced how much support individuals were able to receive from provincial and territorial social assistance.

The experiences of food insecurity reported by adolescents is worrisome, given the documented associations between food insecurity and high healthcare costs, deleterious health outcomes, and poorer academic achievement (Clemens et al., 2024; Faught et al., 2017; Frongillo et al., 2024; Kirkpatrick & Tarasuk, 2008; Ovenell et al., 2022). Child- and adolescent-reported food insecurity, but not adult-reported food insecurity, is associated with school absenteeism, engaging in labour, and lower quality diets (Bernal et al., 2014, 2016). In addition to underscoring the pressing need for policy interventions to ameliorate household food insecurity, these findings point to the importance of interventions, such as facilitating access to mental health support within and beyond school settings, to mitigate negative long-term consequences among adolescents who have experienced food insecurity (Aguirre Velasco et al., 2020; Frongillo et al., 2024).

A strength of this investigation was use of the CFIES, which enabled direct adolescent reporting (Frongillo et al., 2022). The cross-sectional data preclude inferring causality, although the observed correlates of food insecurity have been documented numerous times within the literature (Hutchinson & Tarasuk, 2022; Kirkpatrick & Tarasuk, 2008; Li et al., 2023; Liu et al., 2023). Respondents were recruited using nonprobability-based sampling, and the analyses do not include adolescents living in the territories, where the prevalence of food insecurity among adolescents is likely higher (Statistics Canada, 2023). Provincial and territorial estimates from prior research are not directly comparable to estimates in the present investigation due to differences in the level at which food insecurity is estimated, i.e., household versus person-level, and who reported the food insecurity experiences, i.e., adults versus adolescents (Li et al., 2023). The comparability of the sample composition to the general population of adolescents within the provinces was, however, improved with weighting by age group, sex-at-birth, and region.

## Conclusion

Our understanding of food insecurity among adolescents has been primarily drawn from adult reports of their experiences, resulting in an incomplete understanding of the extent and nature of adolescent food insecurity in Canada. In the current study, *worrying about food scarcity*, *worrying about parental/guardian ability to get food*, and *not being able to get the food they wanted* were most frequently affirmed by adolescents. Close to or more than one in four adolescents had experiences related to the *uncertainty* and *compromised diet quality or preferences* subconstructs. Policy responses with the potential to prevent food insecurity and mitigate its negative consequences among adolescents are needed. Use of the CFIES to monitor adolescent experiences of food insecurity in routine surveillance in Canada, including the three territories, is warranted and can assist with the implementation and evaluation of programs and policies with the potential to reduce the prevalence and number of food insecurity experiences among adolescents, as well as the adverse consequences.

## Contributions to knowledge

What does this study add to existing knowledge?

- Food insecurity among adolescents is typically reported by adults, providing an incomplete and inaccurate understanding of adolescent experiences.
- Around one in three adolescents living in Canada’s provinces reported at least one food insecurity experience in each of 2019, 2020, and 2021, as measured by the Child Food Insecurity Experiences Scale, a direct measure of child and adolescent food insecurity.
- Identifying as Black, East/Southeast Asian, Indigenous, Latino, and South Asian and reporting their family did not have enough money for things they needed were associated with having at least one food insecurity experience in the previous year.

What are the key implications for public interventions, practice or policy?

- Findings reinforce the need for public interventions and policies to address food insecurity among adolescents and to ameliorate the negative implications for those who have experienced it.
- Routine measurement of food insecurity among adolescents using direct assessment methods (e.g., the 10-item Child Food Insecurity Experiences Scale) is warranted and can improve the ability to address food insecurity experiences relevant to this age group by understanding the range of experiences they can have.

## Authorship

The authors’ responsibilities were as follows – AP and SIK: formulated the research questions; AP: conducted the analyses and led the drafting of the manuscript; DH and LV: designed the International Food Policy Study; all authors: provided critical feedback and read and approved the final manuscript.

## Declarations

### Ethics approval

The study was reviewed by and received ethics clearance through a University of Waterloo Research Ethics Board (REB# 41477).

### Consent to participate

Adults who confirmed they had an adolescent aged 10 to 17 years living in their household were asked for permission for their adolescent to complete a web-based survey. Adolescent respondents were asked to provide assent.

### Consent for publication

Not applicable.

### Availability of data and material

Data described in the manuscript, code book, and analytic code can be accessed upon request pending application and approval. Study information can be found on the International Food Policy Study website: http://foodpolicystudy.com.

### Code availability

Analyses were conducted using SAS version 9.4 (Cary, NC).

### Declaration of Generative AI and AI-assisted technologies in the writing process

The authors declare that no generative AI or AI-assisted technologies were used in the writing of this manuscript.

